# Prevalence of hepatitis A virus among migrant workers in Qatar: A national study

**DOI:** 10.1101/2024.02.27.24303440

**Authors:** Nadin Younes, Hiam Chemaitelly, Parveen Banu Nizamuddin, Tasneem Al-Hamad, Marah Abdallah, Hadi M Yassine, Laith J Abu-Raddad, Gheyath K. Nasrallah

**Affiliations:** Department of Biomedical Science, College of Health Sciences, QU Health, Qatar University, Doha, Qatar; Biomedical Research Center, Qatar University, Doha, Qatar; Infectious Disease Epidemiology Group, Weill Cornell Medicine-Qatar, Cornell University, Doha, Qatar; World Health Organization Collaborating Centre for Disease Epidemiology Analytics on HIV/AIDS, Sexually Transmitted Infections, and Viral Hepatitis, Weill Cornell Medicine–Qatar, Cornell University, Qatar Foundation – Education City, Doha, Qatar; Department of Population Health Sciences, Weill Cornell Medicine, Cornell University, New York, New York, USA; Qatar Biobank, Doha, Qatar; Clinical Microbiology & Virology division, Hamad General Hospital, Doha, Qatar; Department of Public Health, College of Health Sciences, QU Health, Qatar University, Doha, Qatar; College of Health and Life Sciences, Hamad bin Khalifa University, Doha, Qatar

**Author notes:** Correspondence Dr. Gheyath K. Nasrallah, Department of Biomedical Science, College of Health Sciences, Qatar University, Doha 2713, Qatar.

**Keywords:** Hepatitis, HAV, prevalence, infection, workers, cross-sectional, Qatar

## Abstract

**Background:** Hepatitis A virus (HAV) is the predominant cause of acute viral hepatitis worldwide; however, data on HAV antibody prevalence (seroprevalence) among migrant populations are limited. This study aimed to investigate HAV seroprevalence among Qatar’s migrant craft and manual workers (CMWs), constituting approximately 60% of the country’s population.

**Methods:** HAV antibody testing was conducted on stored serum specimens obtained from CMWs during a nationwide severe acute respiratory syndrome coronavirus 2 (SARS-CoV-2) population-based cross-sectional survey between July 26 and September 9, 2020. Associations with HAV infection were investigated through regression analyses.

**Results:** Of the 2,607 specimens with HAV antibody test results, 2,393 were positive, and 214 were negative. The estimated HAV seroprevalence among CMWs was 92.0% (95% CI: 90.9- 93.1%). HAV seroprevalence was generally high but exhibited some variation, ranging from 70.9% (95% CI: 62.4-78.2%) among Sri Lankans to 99.8% (95% CI: 98.2-99.9%) among Pakistanis. The multivariable regression analysis identified age, nationality, and educational attainment as statistically significant factors associated with HAV infection. Relative to CMWs aged ≤29 years, the adjusted relative risk (ARR) was 1.06 (95% CI: 1.03-1.10) in CMWs aged 30- 39 years and reached 1.15 (95% CI: 1.10-1.19) in those aged ≥50 years. In comparison to Indians, the ARR was lower among Sri Lankans, assessed at 0.81 (95% CI: 0.72-0.91), but higher among Nepalese at 1.07 (95% CI: 1.04-1.11), Bangladeshis at 1.10 (95% CI: 1.07-1.13), Pakistanis at 1.12 (95% CI: 1.09-1.15), and Egyptians at 1.15 (95% CI: 1.08-1.23). No evidence for differences was found by geographic location or occupation.

**Conclusions:** HAV seroprevalence among Qatar’s CMW population is very high, with over nine out of every ten individuals having been exposed to this infection, likely during childhood.

## INTRODUCTION

Hepatitis A virus (HAV) stands as the predominant cause of acute viral hepatitis globally (1). While HAV often results in mild illness, there are rare instances where the infection progresses to acute liver failure and, ultimately, death (2). Although vaccine preventable (3), the estimated annual incidence of HAV infections exceeds 150 million, with an associated death toll surpassing 39,000 in 2019 (4).

HAV is mainly transmitted via the fecal-oral route through the ingestion of contaminated food and water (5). Other modes of transmission include exposure to infected body fluids or close physical contact such as oral or anal intercourse with an infectious person (6). The largest burden of infection is concentrated in low- and middle-income countries where inadequate sanitary conditions prevail, with most infections occurring asymptomatically during childhood (4). The endemic nature of HAV in these countries leads to frequent exposures in early life and a large proportion of immune adults (7). In higher income countries, susceptible adults face the risk of symptomatic infection, with outbreaks documented among international travelers and specific populations at risk such as men who have sex with men, people who inject drugs, and homeless populations (3, 6). Recent trends also indicate a rise in foodborne outbreaks due to the globalization of trade (7, 8).

The World Health Organization (WHO) formulated the Global Health Sector Strategy 2022-2030, envisioning the elimination of viral hepatitis as a public health concern by 2030 (9). Currently, the WHO recommends universal vaccination against HAV but only in countries where individuals face an intermediate risk of infection, defined as an age-specific prevalence of infection of <90% by 10 years of age but ≥50% by 15 years of age (3, 6, 10). The current approach includes targeted vaccination for high-risk populations in countries with lower infection rates, while routine vaccination is discouraged in endemic countries to avoid paradoxical rise in disease incidence among unvaccinated individuals (10) (3, 6, 10). As of May 2021, immunization against HAV for children had been introduced or was planned to be introduced in only 34 countries (4). Immunization against HAV has been successfully integrated into the national immunization programs of Bahrain, Oman, Qatar, Saudi Arabia, and Tunisia (11). In Iraq, targeted vaccination schemes for high-risk populations are also in place (11). Nevertheless, in most other countries in the region, HAV vaccines are only available at a cost in the private sector (11).

In the Middle East and North Africa (MENA) region, a systematic review of evidence indicated a decline in reported HAV incidence among children over the past two decades (11). However, HAV remains endemic at high incidence in few countries, including Egypt, Morocco, and Pakistan, as well as in geographic areas marked by conflict (11, 12).

Given the limited global data on viral hepatitis infections among migrant populations, this study aimed to determine HAV antibody prevalence (seroprevalence), indicating a history of HAV infection, among Qatar’s craft and manual worker (CMW) population. Qatar, known for its diverse demographics with 89% of the population comprising expatriates from over 150 countries (13), has a large CMW population, constituting approximately 60% of the total population of the country. This population group, primarily consisting of unmarried men aged 20-49 years, is recruited for employment in infrastructure and development projects, including those associated with the World Cup 2022 (14–16). The majority of CMWs originate from countries where HAV infection is endemic at high incidence, and the typical age for acquiring the infection is below 10 years (12, 17). The overarching objective of this study was to provide insights into HAV epidemiology and contribute data to national initiatives aiming to meet global targets for viral hepatitis elimination.

## METHODS

### Study design, sampling, and specimen collection and handling

This study tested deidentified stored blood serum specimens collected during a nationwide serological survey conducted from July 26, 2020, to September 09, 2020 to determine the severe acute respiratory syndrome coronavirus 2 (SARS-CoV-2) seroprevalence among CMWs (14, 18, 19). The survey’s sampling strategy was formulated following an analysis of the registered users’ database of the Qatar Red Crescent Society (QRCS), the primary healthcare provider for CMWs in the country (14).

QRCS manages four centers strategically spread across Qatar to address the healthcare needs of CMWs nationwide (14). These centers cover expansive catchment areas, operate extended hours, and provide services either free of charge or with substantial subsidies (14). To ensure sample representativeness, the probability distribution of CMWs by age and nationality from the QRCS database was cross-referenced with that of expatriate residents from the Ministry of Interior database (20).

With men constituting >99% of CMWs (21), the sampling strategy did not explicitly factor sex. CMWs were recruited at QRCS centers using a systematic sampling approach, informed by the average daily attendance at each center (14). At each center, recruitment proceeded by inviting every 4^th^ attendee to participate in the study until the required sample size was reached for all age and nationality strata. To address challenges in recruiting participants in smaller age-nationality strata, especially among younger individuals of specific nationalities, the recruitment criteria were adjusted towards the conclusion of the study. In these instances, all attendees in these strata were invited to participate, rather than every 4^th^ attendee.

Trained interviewers administered the written informed consent and the study instrument to participants, accommodating their language preferences among nine options: Arabic, Bengali, English, Hindi, Nepali, Sinhala, Tagalog, Tamil, and Urdu (14). The instrument, designed following WHO guidance for developing SARS-CoV-2 sero-epidemiological surveys (22), collected essential socio-demographic information. Certified nurse’s drew a 10 mL blood specimen for serological testing, which was then stored in an icebox before transportation to the Qatar Biobank for long-term storage and subsequent testing.

### Laboratory methods

Serum aliquots were extracted from the archived specimens at the Qatar Biobank (QBB) and subsequently transferred to the virology laboratory at Qatar University (QU) for serological testing. The sera at both the QBB and QU were maintained at -80°C until employed for serology testing.

HAV seroprevalence was determined by testing sera for HAV antibodies using Dia.Pro competitive enzyme linked Immunosorbent Assay (ELISA) From Diagnostic Bioprobes Srl, Milano, Italy (23). This assay has a reported sensitivity of 100% and a specificity exceeding 98% (23). Interpretation of test results followed the manufacturer’s guidelines. Specimens with a mean cut-off/optical density (OD) value at 450nm (designated as cut-off index values) below 0.9 were considered negative, those surpassing 1.1 were considered positive, and those within the range of 0.9 to 1.1 were considered equivocal (23). Equivocal samples were excluded from further analysis.

The mean cut-off value was calculated using the equation: mean cut-off value = (OD value for the negative control well + OD value for the positive control well)/3 (23).

### Oversight

This study received approval from the Institutional Review Boards of Hamad Medical Corporation, Qatar University, and Weill Cornell Medicine-Qatar. The reporting of the study adhered to the Strengthening the Reporting of Observational Studies in Epidemiology (STROBE) guidelines, as detailed in Table S1 in the Online Supplementary Document.

### Statistical analysis

Frequency distributions and measures of central tendency were used to describe study participants. HAV seroprevalence was estimated after applying probability weights to correct for participants’ unequal selection and ensure the sample’s representativeness of the broader CMW population. Weights were calculated using the population distribution of CMWs by age, nationality, and QRCS center, extracted from the QRCS registered-user database (14). HAV cut-off index values obtained from testing with the Dia.Pro competitive ELISA were illustrated using a histogram.

Associations with HAV infection were explored through Chi-square tests and univariable and multivariable Poisson regression analyses with robust error variance, not logistic regression analyses, given the high observed HAV seroprevalence levels (24, 25). Variables with a p-value ≤0.2 in the univariable analyses were included in the multivariable model. A p-value <0.05 in the multivariable analysis was considered indicative of a statistically significant association. Unadjusted and adjusted relative risks (RRs and ARRs, respectively), along with their respective 95% confidence intervals (CIs) and p-values, were reported. Interactions were not considered. All statistical analyses were conducted using Stata/SE version 18.0 (Stata Corporation, College Station, TX, USA).

## RESULTS

### Study population

Table 1 outlines the characteristics of the study participants. Among the 2,641 blood specimens collected during the SARS-CoV-2 seroprevalence survey (14), only 2,616 specimens were available and had sufficient sera for HAV testing. Of these, 2,393 specimens were positive, 214 were negative, and 9 were equivocal and therefore excluded from further analysis.

**Table 1.** Characteristics and seroprevalence of HAV infection among study participants.

| Characteristics | Tested | HAV seroprevalence |  |  |
| --- | --- | --- | --- | --- |
|  | N (%)<sup>*</sup> | N | %<sup>†</sup> (95% CI)<sup>‡</sup> | Chi-square p-value |
| Age (years) |  |  |  |  |
| ≤29 | 747 (27.7) | 637 | 86.4 (83.7-88.7) | <0.001 |
| 30-39 | 966 (41.8) | 893 | 92.5 (90.6-94.0) |  |
| 40-49 | 545 (21.4) | 521 | 96.2 (94.2-97.5) |  |
| 50-59 | 258 (7.4) | 251 | 96.8 (93.3-98.5) |  |
| 60+ | 91 (1.7) | 91 | 100.0 (96.0-100.0)<sup>‡</sup> |  |
| Nationality |  |  |  |  |
| All other nationalities<sup>§</sup> | 231 (7.6) | 210 | 90.6 (85.9-93.8) | <0.001 |
| Bangladeshi | 623 (26.0) | 610 | 97.9 (96.4-98.8) |  |
| Egyptian | 90 (3.2) | 86 | 95.7 (88.2-98.5) |  |
| Filipino | 102 (2.7) | 78 | 78.3 (68.1-86.0) |  |
| Indian | 716 (29.2) | 628 | 87.6 (84.9-89.9) |  |
| Nepalese | 564 (21.7) | 539 | 95.6 (93.5-97.1) |  |
| Pakistani | 136 (4.9) | 135 | 99.8 (98.2-99.9) |  |
| Sri Lankan | 145 (4.8) | 107 | 70.9 (62.4-78.2) |  |
| QRCS center (catchment area within Qatar) |  |  |  |  |
| Fereej Abdel Aziz (Doha-East) | 611 (23.5) | 548 | 89.8 (87.1-92.0) | 0.132 |
| Zekreet (North-West) | 234 (2.3) | 215 | 92.0 (87.7-94.9) |  |
| Hemaila (South-West; "Industrial Area") | 964 (41.9) | 897 | 93.5 (91.8-94.9) |  |
| Mesaimeer (Doha-South) | 798 (32.4) | 733 | 91.8 (89.6-93.5) |  |
| Educational attainment |  |  |  |  |
| Primary or lower | 626 (24.6) | 592 | 94.4 (92.2-96.1) | <0.001 |
| Intermediate | 429 (17.9) | 417 | 97.4 (95.3-98.5) |  |
| Secondary/High school/Vocational | 1,085 (44.1) | 1,003 | 92.7 (91.0-94.2) |  |
| University | 369 (13.4) | 285 | 76.5 (71.6-80.8) |  |
| Occupation |  |  |  |  |
| Professional workers<sup>¶</sup> | 136 (4.9) | 107 | 76.9 (68.4-83.6) | <0.001 |
| Food & beverage workers | 91 (3.2) | 79 | 89.0 (80.7-94.0) |  |
| Administration workers | 82 (3.0) | 73 | 87.5 (77.2-93.5) |  |
| Retail workers | 169 (6.6) | 149 | 90.1 (84.6-93.8) |  |
| Transport workers | 430 (16.4) | 402 | 93.5 (90.6-95.6) |  |
| Security workers | 60 (2.3) | 55 | 91.1 (80.2-96.3) |  |
| Cleaning workers | 105 (4.0) | 96 | 91.7 (84.7-95.6) |  |
| Technical and construction workers<sup>‡</sup> | 1,311 (52.8) | 1,237 | 94.6 (93.1-95.7) |  |
| Other workers<sup>**</sup> | 174 (6.8) | 151 | 86.5 (80.4-90.9) |  |
| **Total (% , 95% CI)** | **2,607 (100.0)** | **2,393** | **92.0 (90.9-93.1)** | **--** |

CI, confidence interval; QRCS, Qatar Red Crescent Society.
<sup>\*</sup>Percentage of the sample weighted by age, nationality, and QRCS center. Missing values for socio-demographic variables were excluded from the analysis.
<sup>†</sup>Percentage of positive out of the total sample weighted by age, nationality, and QRCS center.
<sup>‡</sup>The 95% confidence intervals were calculated using the binomial distribution.
<sup>§</sup>Includes all other nationalities of craft and manual workers residing in Qatar.
<sup>¶</sup>Includes architects, designers, engineers, operation managers, and supervisors among other professions.
<sup>‡</sup>Includes carpenters, construction workers, crane operators, electricians, foremen, maintenance/air conditioning/cable technicians, masons, mechanics, painters, pipe-fitters, plumbers, and welders among other professions.
<sup>\*\*</sup>Includes barbers, firefighters, gardeners, farmers, fishermen, and physical fitness trainers among other professions.

Around 70% of the study participants were younger than 40 years, with a median age of 35.0 years and an interquartile range (IQR) spanning 29.0-43.0 years. Slightly more than 40% of participants had educational attainment at intermediate or lower levels, 44.1% had high school education or vocational training, and 13.4% had higher educational attainment. The distribution of nationalities highlighted a prevalent presence of Indians (29.2%), Bangladeshis (26.0%), and Nepalese (21.7%), aligning with the broader demographic composition of the CMW population in Qatar (20). Over half of CMWs (52.8%) were employed in technical and construction jobs, which included occupations such as carpenters, crane operators, electricians, masons, mechanics, painters, plumbers, and welders.

### HAV seroprevalence and associations with infection

HAV seroprevalence among CMWs was estimated at 92.0% (95% CI: 90.9-93.1%). HAV cut-off index values in positive specimens ranged from 1.1 to 485.0 (Figure 1), with a median of 22.8 (IQR: 15.9-27.3).

**Fig 1.**
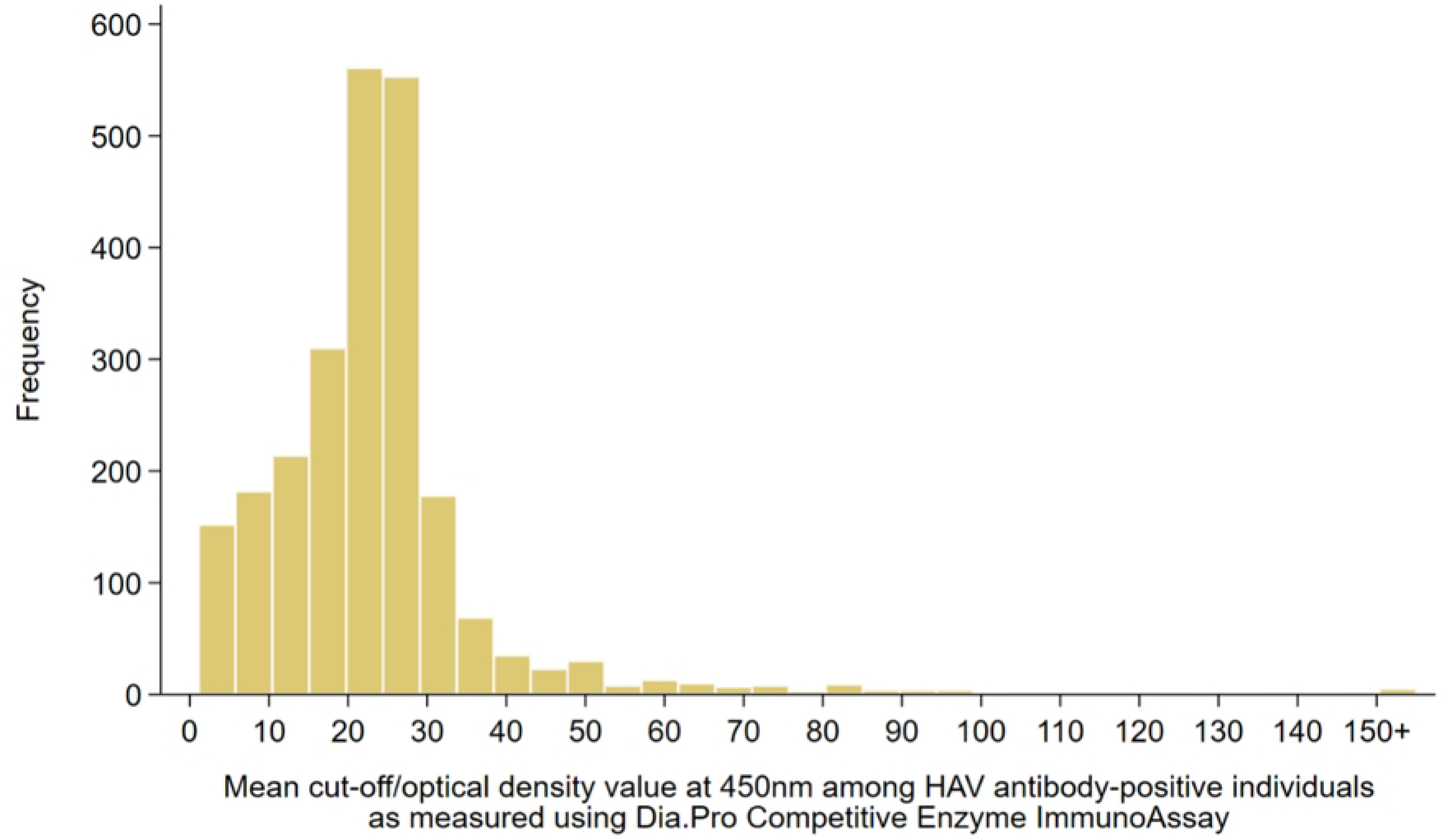
Distribution of the mean cut-off/optical density (OD) values at 450nm among HAV antibody-positive individuals as measured using the Dia.Pro Competitive ELISA.

Table 1 shows HAV seroprevalence estimates across various socio-demographic characteristics within the population. Figure 2 illustrates HAV seroprevalence by nationality group. Despite variations, HAV seroprevalence was generally high ranging from 70.9% (95% CI: 62.4-78.2%) among Sri Lankans to 99.8% (95% CI: 98.2-99.9%) among Pakistanis.

**Fig 2.**
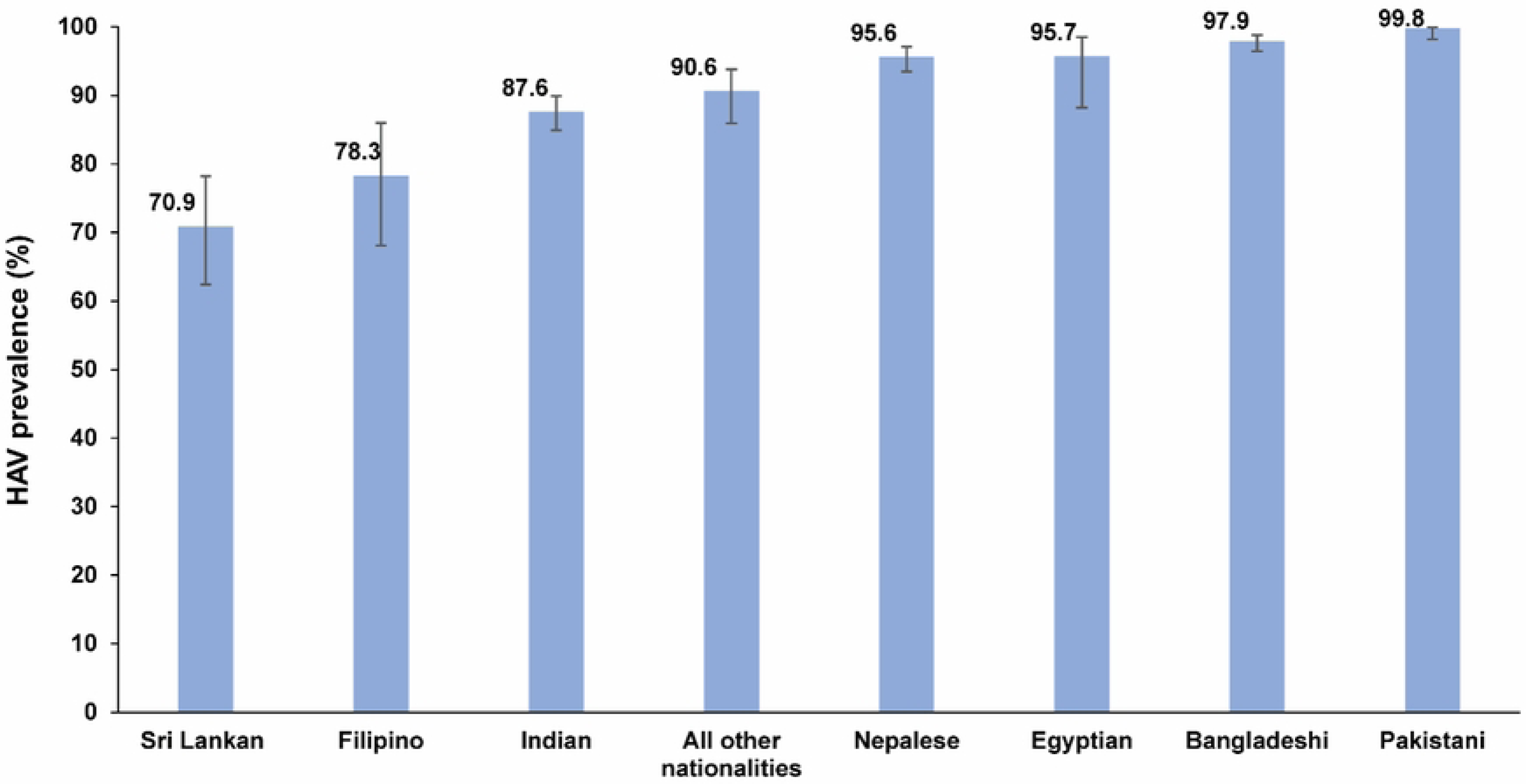
HAV seroprevalence by nationality group among the craft and manual worker population in Qatar.

The multivariable regression analysis identified age, nationality, and educational attainment as statistically significant factors associated with HAV infection (Table 2). The ARR increased gradually with age. Relative to CMWs ≤29 years of age, the ARR was 1.06 (95% CI: 1.03-1.10) in CMWs aged 30-39 years and reached 1.15 (95% CI: 1.10-1.19) in those aged ≥50 years. There were significant differences by nationality, where compared to Indians, the ARR was lower among Sri Lankans assessed at 0.81 (95% CI: 0.72-0.91), but higher among Nepalese at 1.07 (95% CI: 1.04-1.11), Bangladeshis at 1.10 (95% CI: 1.07-1.13), Pakistanis at 1.12 (95% CI: 1.09-1.15), and Egyptians at 1.15 (95% CI: 1.08-1.23).

**Table 2.**
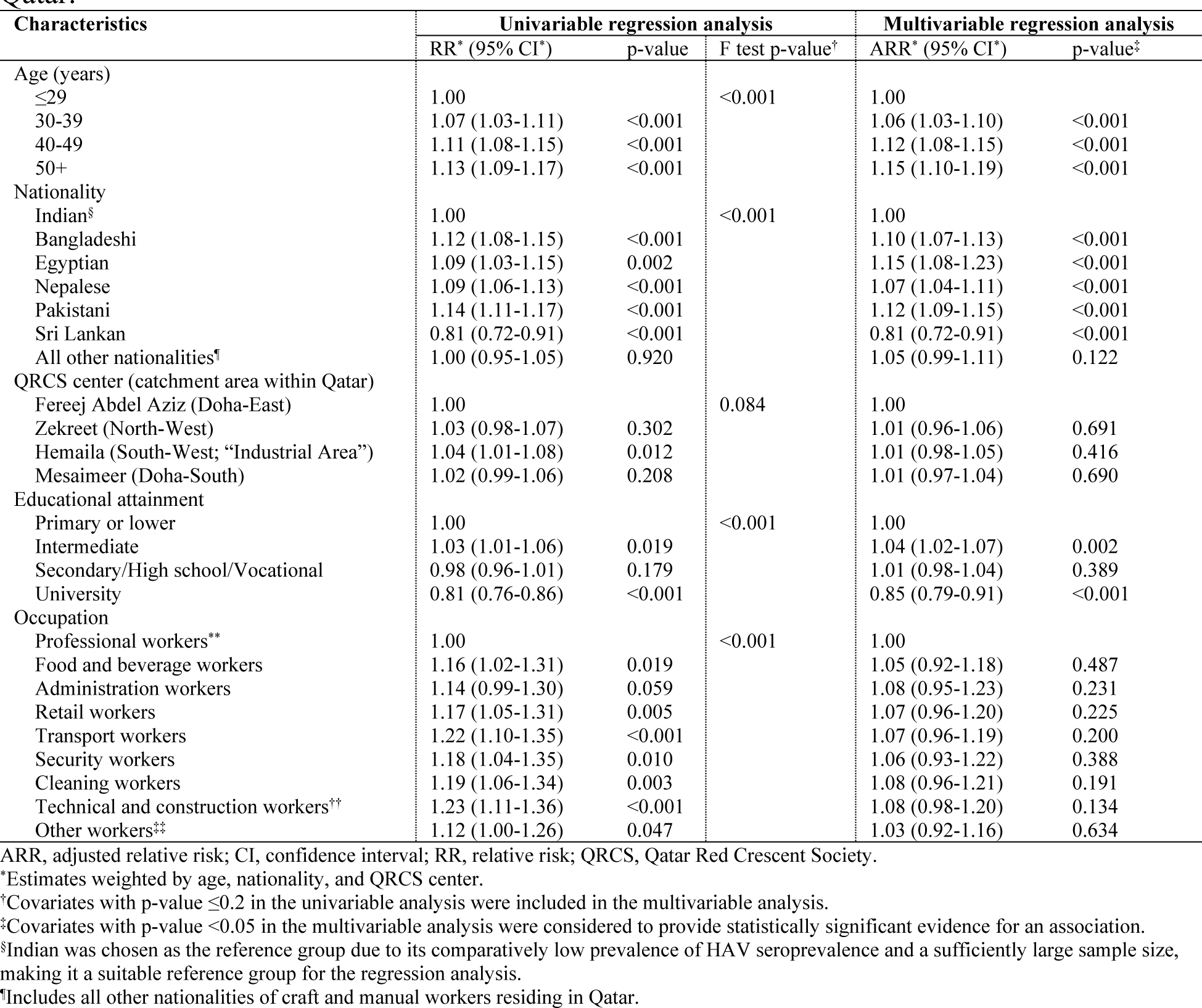
Associations with HAV infection among the craft and manual worker population in Qatar.

Significant differences were observed based on educational attainment. CMWs with intermediate education had an ARR of 1.04 (95% CI: 1.02-1.07) while those with university education had an ARR of 0.85 (95% CI: 0.79-0.91), both compared to those with primary or lower education. No evidence for differences was found by QRCS center (proxy of catchment area/geographic location) or by occupation.

## DISCUSSION

HAV seroprevalence is highly elevated among the CMW population in Qatar, with over nine out of every ten individuals having experienced this infection. This observation suggests that nearly the entire CMW population has acquired the infection, predominantly during childhood before relocating to Qatar, considering the very high HAV seroprevalence observed in the migrants’ countries of origin (12, 17). This underscores the significance of a comprehensive plan for HAV prevention. Such a plan should incorporate measures to promote safe water and sanitation practices, along with adherence to WHO recommendations for HAV vaccination in countries where individuals face a heightened risk of symptomatic infection (3, 6, 10).

While HAV seroprevalence was generally very high, some variations were observed. These variations included differences by nationality, with Sri Lankans exhibiting the lowest seroprevalence and Nepalese, Bangladeshis, Pakistanis, and Egyptians having the highest seroprevalence. Seroprevalence was at its lowest among individuals with a university education, suggesting an association between infection and lower socio-economic status, as observed elsewhere (17). Seroprevalence increased with age, consistent with expectations for a measure of cumulative exposure to the infection. However, the rate of increase with age was slow, indicating that only a minority of infections are acquired in adulthood.

Our study represents the first endeavor to estimate the seroprevalence of HAV in Qatar, filling a critical gap in the existing research. Its significance is underscored by the scarcity of studies on HAV seroprevalence in the Middle East, particularly among QRCS and the Gulf region, where CMWs constitute a significant demographic. Prominent strengths of our study include a substantial sample size and the utilization of a highly sensitive ELISA for detecting HAV antibodies, enhancing the reliability and accuracy of the seroprevalence results.

This study is subject to certain limitations. Originally designed to employ a probability-based sampling strategy for CMW recruitment, logistical challenges led to the implementation of a systematic sampling method, concentrating on QRCS attendees. In response to this modification, probability-based weights were introduced to produce a seroprevalence estimate that aligns with the broader CMW population. To account for the representation of smaller age-nationality strata, a deviation from the initial plan—selecting every fourth attendee—was introduced. Towards the end of the study, all individuals in these strata were approached to participate, ensuring a more comprehensive representation of the sampled population.

Operational challenges presented difficulties in monitoring and maintaining consistent records of the response rate among nurses in the QRCS centers. As a result, an exact estimate of the response rate could not be determined, but it was approximated to exceed 90% based on the interviewers’ experience. While there is potential for the recruitment scheme to impact the generalizability of the study findings, this is considered less likely given the substantial daily influx of CMWs visiting these centers, surpassing 5,000 patients daily (14). Of note that these centers serve as the primary healthcare providers for CMWs in the country, offering a range of services beyond patient treatment, including periodic health certifications, vaccinations, and pre-travel SARS-CoV-2 testing.

## CONCLUSIONS

This study revealed a very high HAV seroprevalence among Qatar’s CMW population, with more than 90% having experienced the infection. This underscores that virtually the entire CMW population contracted the infection, presumably during childhood before their migration to Qatar, given the comparably elevated HAV seroprevalence in their countries of origin. Despite the overall elevated seroprevalence, notable variations were observed, particularly across nationalities and socio-economic strata. These findings contribute considerations for healthcare service planning and policy development. This aims to alleviate the burden of HAV-related illnesses and work towards the 2030 targets for viral hepatitis elimination.

## Acknowledgements

We thank all participants for their willingness to be part of this study. We thank all members of the Craft and Manual Workers Seroprevalence Study Group for their efforts in supporting the collection of the samples of this study. We also thank Dr. Nahla Afifi, Director of Qatar Biobank (QBB), Ms. Tasneem Al-Hamad, Ms. Eiman Al-Khayat, and the rest of the QBB team for their unwavering support in retrieving and analyzing samples. We also acknowledge the dedicated efforts of the Surveillance Team at the Ministry of Public Health for their support in sample collection.

## Authorship contributions

GKN conceived the study and led the laboratory testing. NY, PBN, TAH, MA, and HMY conducted the lab testing of the specimens. GKN and NY validated and interpreted the laboratory results. HC developed the study design, managed the databases, performed the data analyses, and wrote the first draft of the article. LJA led the statistical analyses and contributed to the first draft of the article. GKN and LJA critically reviewed the manuscript. All authors have read and approved the final manuscript.

## Data availability statement

All data are available in aggregate form within the manuscript.

## Ethics approval

Hamad Medical Corporation, Qatar University (QU-IRB 1558-EA/21), and Weill Cornell Medicine-Qatar Institutional Review Boards approved this study.

